# Natural History Of Oral Human Papillomavirus In Nigerian Mothers And Youth With And Without HIV: A Prospective Cohort Study

**DOI:** 10.64898/2026.11.05.26367373

**Authors:** Anil Kumar, Oluwaseun Peter, Esosa Osagie, Jia Liu, Morgan C. Byrd, Jianhong Chen, Paul Akhigbe, Nosakhare Lawrence Idemudia, Ibukun J Abulude, Peter Kavecky, Ozoemene Obuekwe, Fidelis Ewenitie Eki-Udoko, Nicolas F Schlecht, Yana Bromberg, Nosayaba Osazuwa-Peters, Modupe O Coker

**Author notes:** Corresponding Author: Dr. Modupe O Coker Associate Professor, Basic and Translational Sciences and Epidemiology, Center for Clinical and Translational Research, School of Dental Medicine, University of Pennsylvania Philadelphia, Pennsylvania 19104.

## Abstract

**Objectives:** HPV-associated oropharyngeal cancer is rising, yet the natural history of oral-HPV, particularly persistence remains poorly characterized in sub-Saharan Africa and in people living with HIV (PLWH). We characterized oral HPV prevalence, incident detection, persistence, and clearance over four visits in a Nigerian mother-youth cohort with and without HIV and quantified the genotype distribution relative to vaccine coverage.

**Methods:** Within the prospective HOMINY cohort, 1075 participants (404 mothers and 671 youth) were followed across four visits at 6-month intervals. Oral rinses were genotyped with the Seegene Allplex HPV28 assay (28 genotypes). Persistence was defined as detection at ≥2 visit (not necessarily consecutive), with type-specific and visit 2-4 sensitivity definitions. We used modified-Poisson models for visit-level detection, Markov multistate models for acquisition and clearance, and a permutation test for mother-youth genotype concordance.

**Results:** Oral HPV was detected at least once in 112 participants (10.4% overall; 11.1% mothers and 10% youth); incident detection was around 6% in both groups. Persistence was uncommon (1.9%), and only 3 had oral HPV DNA detected at all four visits. Clearance predominated across every visit interval. In Markov models, clearance was slower in PLWH (HR 0.42, 95% CI 0.22-0.77) with no difference in acquisition (HR 0.77, 95% CI 0.39-1.5). Persistence concentrated among PLWH. Only 31.2% of ever-positive participants carried a Gardasil-9 preventable strain of HPV; HPV16 and HPV39 were most frequent. Mother-youth genotype concordance exceeded chance (permutation p =0.009).

**Conclusion:** Oral HPV was common but transient; persistence was confined to a small PLWH minority and was dominated by high-risk non-vaccine genotypes, leaving most infections beyond the reach of current vaccines.

## Introduction

Human papillomavirus (HPV) infection is among the most common sexually transmitted infections worldwide and contributes to approximately one in three cancers, including cancers of the cervix and oropharynx.^1,2^ Although cancers of the oral cavity have declined over the past two decades, the incidence of HPV-associated oropharyngeal cancer (OPC) has risen sharply^3^, now surpassing that of HPV-related cervical cancers, and is projected to keep rising over the next 30 to 40 years.^4^ It is important to understand the dynamics of oral HPV persistence and clearance,^5^ since most infections clear spontaneously within two years^2^ while a small persistent infections carry oncogenic risk.^6^ Persistence is difficult to study because there is no universal definition, and the host factors that distinguish persistent from transient infection are not fully characterized.^6^

The genotypes that persist also vary by region. Globally, HPV16 dominates oral infection and HPV-attributable OPCs, and is the oncogenic type least likely to clear^7^, but the pooled International Agency for Research on Cancer surveys identified Nigeria among the countries with the highest HPV prevalence worldwide, where women with HPV are more likely to carry other high-risk genotypes like HPV35, HPV45, HPV52, HPV56, and HPV58.^8^ HPV35 is disproportionately common across West Africa, contributing a far larger share of high-grade cervical disease and invasive cancer than its roughly 2% global contribution would predict.^9^ Immunosuppression in people living with HIV (PLWH) also broadens the genotype spectrum, favoring infection with multiple strains and with genotypes other than HPV16.^10,11^ Among PLWH in Africa, the genotypes most likely to persist may therefore differ from those that drive OPC in better-studied settings with PLWH. Because a precursor’s oncogenic potential is genotype-specific, documenting which genotypes persist locally bears directly on both risk stratification and the reach of current vaccines.

These dynamics matter most where they are least studied. Sub-Saharan Africa carries the largest share of the global HIV epidemic.^12^ The expansion of antiretroviral therapy (ART) has produced a growing generation of PLWH and children with perinatally exposure to HIV,^13,14^ and their oral HPV experience is poorly understood. Chronic immunosuppression alters HPV natural history at every stage, causing higher prevalence, faster acquisition, slower clearance, and greater persistence among PLWH, with lower CD4 counts and detectable viral loads independently predicting persistent infection.^15^ For oral HPV the pattern is described inadequately, though PLWH cohorts confirms that although most oral infections clear within two years, oncogenic persistence is meaningfully elevated and HPV16 in particular is significantly less likely to clear than other high-risk types.^7^ Higher oral HPV16 viral load has shown to predict long-term persistence in individuals with or at risk for HIV^16^, and the incidence of high-risk oral HPV has been reported several-fold higher in men with HIV than in those without.^17^ Nigeria, with roughly two million PLWH^18^, is a setting in which these interactions are consequential but largely uncharacterized.

Nigerian baseline data reflect a correspondingly wide type spectrum, with the low-risk type HPV81 among the most frequently detected across groups.^18^ Oral HPV in children and adolescents is uncommon, and its routes of acquisition; vertical transmission during pregnancy or delivery or horizontal contact within the household remain debated.^19,20^ Prospective family cohorts have demonstrated genotype-specific between maternal-infant concordance at birth consistent with vertical transmission^19^, but also that some persistent childhood oral infections are acquired horizontally from the mother after birth, indicating that mother-to-child transmission is an ongoing rather than a purely peripartum process.^20^ Sampling mothers together with their children who share a household, a behavioral environment, and, frequently, an HIV context allows genotype concordance within a dyad to signal shared or transmitted infection in a way that sampling individuals in isolation cannot, while situating adolescent infection within the maternal reservoir from which much of it may derive.

Ultimately, identifying which genotypes circulate and persist matters because prevention depends on a match between vaccine composition and those genotypes. The nonavalent vaccine (Gardasil-9), a formulation built around the genotypes that dominate cervical disease globally, yet several genotypes common in West Africa like HPV35 falls outside its coverage.^9^ In sub-Saharan Africa this excluded fraction is not trivial; comprehensive genotyping of cervical carcinomas in Malawi estimated that the nonavalent vaccine would prevent roughly 71% of cases, with HPV35 identified as the principal reason for the residual gap.^21^ Nigeria’s national program compounds this ceiling, having introduced the single-dose quadrivalent vaccine (Gardasil-4) into routine immunization for girls aged 9-14 years in October 2023; a choice justified by the lower cost and simpler logistics of a single-dose schedule in a resource-limited setting, but one that covers fewer high-risk types than the nonavalent formulation.^22,23^

Against this background, we leveraged four rounds of longitudinal follow-up in a Nigerian mother-youth cohort to characterize the natural history of oral HPV. We estimated visit-specific positivity and incident detection, quantified persistence and clearance, and assessed the fraction of circulating and persisting types preventable by the nonavalent vaccine.

## Methods

### Study design and population

This analysis used longitudinal data from the Human Papillomavirus, Human Immunodeficiency Virus, and Oral Microbiota Interplay in Nigerian Youth (HOMINY) study^24,25^, a prospective cohort at the University of Benin Teaching Hospital, Nigeria. Between October 2022 and May 2023, youth aged 9-18 years and their mothers were enrolled and subsequently followed across four study visits. Youths were classified as HIV-infected (HI), HIV-exposed uninfected (HEU), or HIV-unexposed uninfected (HUU), and mothers as HIV-infected (HI) or uninfected (HU); youths in the HI and HEU groups were age- and sex-matched to those in the HUU group, which served as the reference for all comparisons.

Recruitment was carried out by trained healthcare professionals who approached eligible mother-youth dyads during routine clinic visits, enrolling consecutively presenting pairs. Written informed consent was obtained from a parent or legal guardian for each minor, and written assent from each youth, before recruitment. Additional verbal consent was obtained for blood and oral sample collection beyond the written consent and assent provided at the visit. Research staff delivered standardized written and verbal explanations of study procedures, risks, and benefits, and individuals who expressed interest and met eligibility criteria were enrolled and retained for longitudinal follow-up. The institutional review boards at Rutgers State University (Pro2022000949) and the UBTH, Benin City (ADM/E22/A/VOL. VII/14813674), Nigeria, approved this study and will be informed of the study findings in a timely manner. University of Pennsylvania and Duke University relied on Rutgers and UBTH for IRB oversight. All procedures involving human participants were conducted in accordance with relevant ethical guidelines and regulations, including the Declaration of Helsinki and its later amendments. To account for an anticipated 15% loss to follow-up, targeted enrollment was 230 youth-mother pairs in each group (n=690), stratified by HIV exposure and infection status, with HUU participants serving as the reference for comparisons. Assuming an attrition-adjusted cohort of approximately 600 participants, we estimated 80% power to detect incidence rate ratios (IRR) of approximately 1.19 or higher via Poisson regression (α=0.05, two-sided). Power was expected to remain adequate for IRRs of approximately 1.20 after adjustment for additional covariates.

### Sample and Data collection

Standardized questionnaires captured sociodemographic, behavioral, and sexual-health variables, and WHO-protocol oral examinations recorded the DMFT, PUFA, OHIS, and HIS indices; full instruments are detailed in the cohort protocol.^24,25^ Whole blood was collected for CD4 and CD8 counts and, where available, HIV viral load. At each visit, participants provided a 10mL saline oral rinse, which was centrifuged at 2000g for 15mins to pellet and resuspended in 4.5mL of the rinse, aliquoted and stored at – 80°C.^24^

### HPV testing and genotyping

Dual HPV testing was performed at laboratories in Nigeria (Inqaba Biotechnical Industries) and New York, USA (Roswell Park Comprehensive Cancer Center) using an identical protocol. DNA was extracted from oral rinse samples with the Zymo Quick-DNA Miniprep Kit and tested with the Seegene Allplex HPV28 assay, which detects 19 high-risk and 9 low-risk genotypes. A participant was classified as positive at a given visit when HPV DNA was detected in that visit’s oral rinse sample. Following established international (IARC) criteria, high-risk genotypes were HPV16, 18, 31, 33, 35, 39, 45, 51, 52, 56, 58, and 59, (with probable high-risk 26, 53, and 66); all other detected genotypes (e.g., HPV6, 11, 40, 42, 43, 44, 54, 61, 68, 73, 70, 72, 82, 81) were classified low risk.^26^

### Retention and handling of missing visits

Visit attendance was determined from recorded visit dates and confirmed using internal-control signals from the assay. No participants returned after missing a scheduled visit, resulting in monotone dropout, with each participant’s follow-up characterized by their last observed visit. This structure informed the handling of infection clearance. Participants whose last observed result was positive were censored at their last observed visit and were not classified as having cleared the infection, since clearance could not be confirmed.

### HPV Persistence Definitions

Visit-specific positivity was expressed using denominators restricted to participants observed at that visit. Incident acquisition was defined as the first detection of oral HPV among participants who were negative at baseline. The primary definition of persistence was detection of oral HPV at two or more of the four visits of any type and not necessarily at consecutive visits.^27^ We examined three alternative definitions: type-specific persistence, requiring the same genotype at two or more visits; and, for both the any-type and type-specific definitions, a window restricted to visits 2 through 4. The visit 2-4 window served as a sensitivity analysis motivated by the possibility of sample contamination or non-standardized handling at baseline. Clearance was defined as a negative result at a later observed visit following detection.

In addition to the any-type and type-specific definitions, we classified each participant into one of four mutually exclusive, hierarchical categories; persistently negative (never positive), transient (positive at a single visit; both high-risk and low-risk), persistent low-risk only (≥2 positive visits, low-risk genotypes only), and persistent high-risk (≥2 positive visits with any high-risk genotype) assigning each participant to the highest applicable category (high-risk > low-risk > transient > negative).

### Statistical analysis

Positivity, acquisition, persistence, and transition frequencies were summarized descriptively; continuous variables are reported as mean (standard deviation) or medians (interquartile range), and categorical variables as counts and percentages. The four-category persistence classification was compared by maternal HIV status and youth HIV exposure group, separately using Fisher’s exact test. The primary outcome was visit-level detection of any high-risk oral HPV, with any-type detection a secondary outcome; we fit Poisson models with family-clustered robust standard errors to estimate prevalence rate ratios (PRs), separately for mothers and youth, with an exploratory generation x HIV interaction model. To characterize HPV acquisition and clearance, we fit a participant-level two-state Markov multistate model with HIV status as a covariate and estimated hazard ratios (HRs) for transition into HPV-positive and HPV-negative states. To assess whether mother-youth dyads shared genotypes more than expected by chance, we compared the observed number of genotype-concordant dyads with a null distribution from 10,000 random mother-youth re-pairings. Analyses used R, with two-sided tests at α=0.05.

## Results

Of the 1,075 enrolled participants (404 mothers and 671 youth), 920 (85.6%) completed all four visits, with monotone attrition and no re-entry after a missed visit **(Figure 1)**. Oral HPV was detected at least once in 112 participants (10.4%), a frequency that was nearly identical in mothers (45; 11.1%) and youth (67; 10%). Visit-specific prevalence was highest at baseline (mothers 5.2% [21/404]; youth 4% [27/671]) and, rather than declining monotonically, followed a shallow U-shaped course, falling to its lowest point at visits 2 and 3 before rebounding at visit 4 (mothers 3.8%; youth 4.1%) **(Figure 2, Table 1)**. This rebound is consistent with continued incident acquisition offsetting clearance among participants negative at baseline, incident detection occurred in 24 of 383 mothers (6.3%) and 40 of 644 youth (6.2%), again virtually identical between cohorts so that new infections largely replaced those that cleared.

**Table 1:** Visit-specific oral HPV positivity, with denominators restricted to participants observed at each visit.

| **Visit** | **Mothers, positive/observed (%)** | **Youth, positive/observed (%)** |
|---|---|---|
| 1 | 21/404 (5.2%) | 27/671 (4.0%) |
| 2 | 14/385 (3.6%) | 17/636 (2.7%) |
| 3 | 12/359 (3.3%) | 20/597 (3.4%) |
| 4 | 13/340 (3.8%) | 24/580 (4.1%) |

**Table 2:**
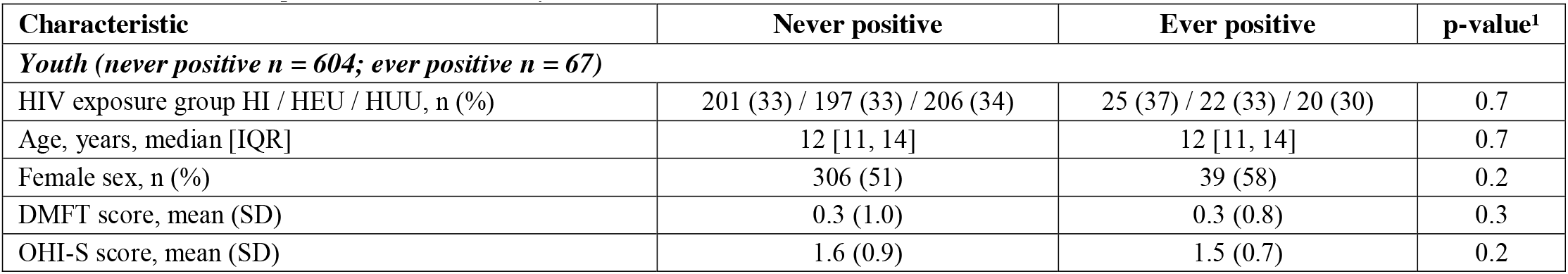
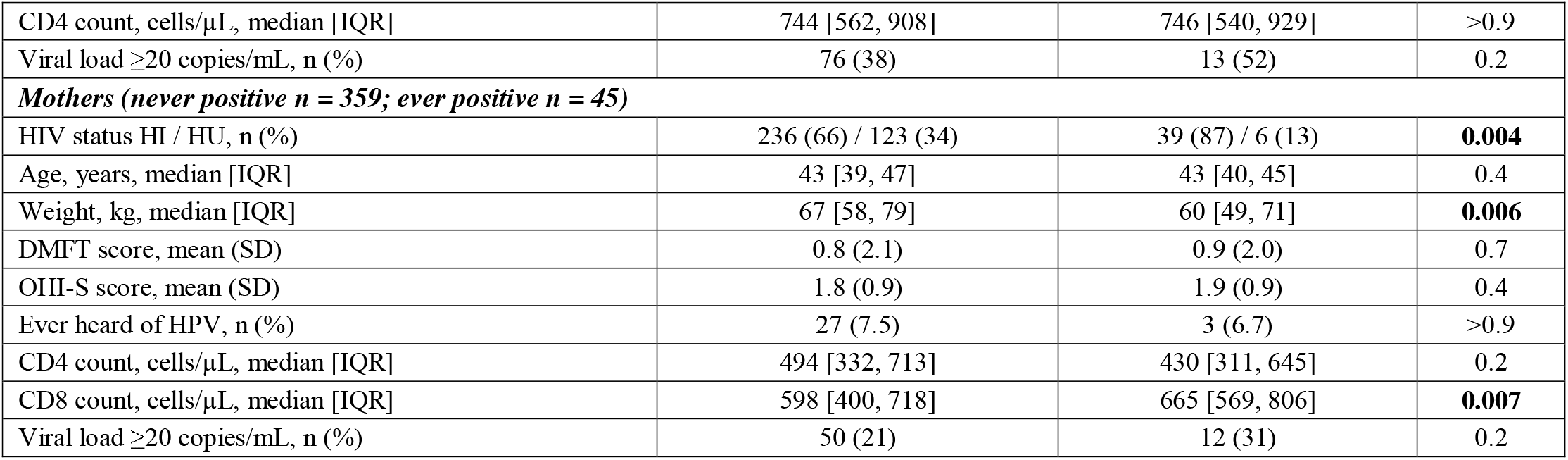
Participant characteristics by cohort and oral HPV status.

| Characteristic | Never positive | Ever positive | p-value<sup>1</sup> |
|---|---|---|---|
| ***Youth (never positive n = 604; ever positive n = 67)*** |  |  |  |
| HIV exposure group HI / HEU / HUU, n (%) | 201 (33) / 197 (33) / 206 (34) | 25 (37) / 22 (33) / 20 (30) | 0.7 |
| Age, years, median [IQR] | 12 [11, 14] | 12 [11, 14] | 0.7 |
| Female sex, n (%) | 306 (51) | 39 (58) | 0.2 |
| DMFT score, mean (SD) | 0.3 (1.0) | 0.3 (0.8) | 0.3 |
| OHI-S score, mean (SD) | 1.6 (0.9) | 1.5 (0.7) | 0.2 |
| CD4 count, cells/µL, median [IQR] | 744 [562, 908] | 746 [540, 929] | >0.9 |
| Viral load ≥20 copies/mL, n (%) | 76 (38) | 13 (52) | 0.2 |
| ***Mothers (never positive n = 359; ever positive n = 45)*** |  |  |  |
| HIV status HI / HU, n (%) | 236 (66) / 123 (34) | 39 (87) / 6 (13) | **0.004** |
| Age, years, median [IQR] | 43 [39, 47] | 43 [40, 45] | 0.4 |
| Weight, kg, median [IQR] | 67 [58, 79] | 60 [49, 71] | **0.006** |
| DMFT score, mean (SD) | 0.8 (2.1) | 0.9 (2.0) | 0.7 |
| OHI-S score, mean (SD) | 1.8 (0.9) | 1.9 (0.9) | 0.4 |
| Ever heard of HPV, n (%) | 27 (7.5) | 3 (6.7) | >0.9 |
| CD4 count, cells/µL, median [IQR] | 494 [332, 713] | 430 [311, 645] | 0.2 |
| CD8 count, cells/µL, median [IQR] | 598 [400, 718] | 665 [569, 806] | **0.007** |
| Viral load ≥20 copies/mL, n (%) | 50 (21) | 12 (31) | 0.2 |

**Table 2:**
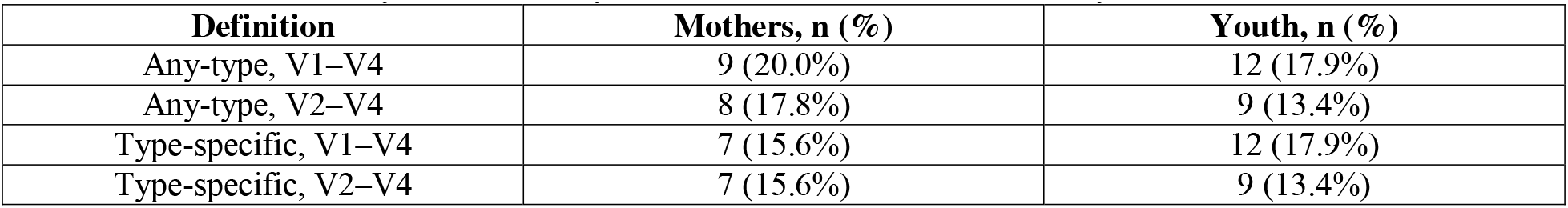
Persistence under four analytic definitions, expressed as a percentage of ever-positive participants.

**Table 3.**
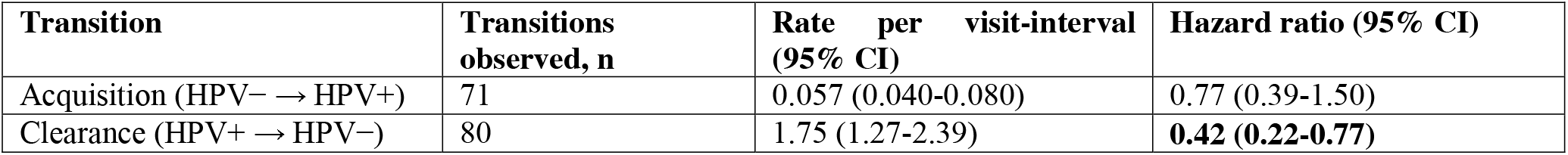
Markov Model.

**Figure 1:**
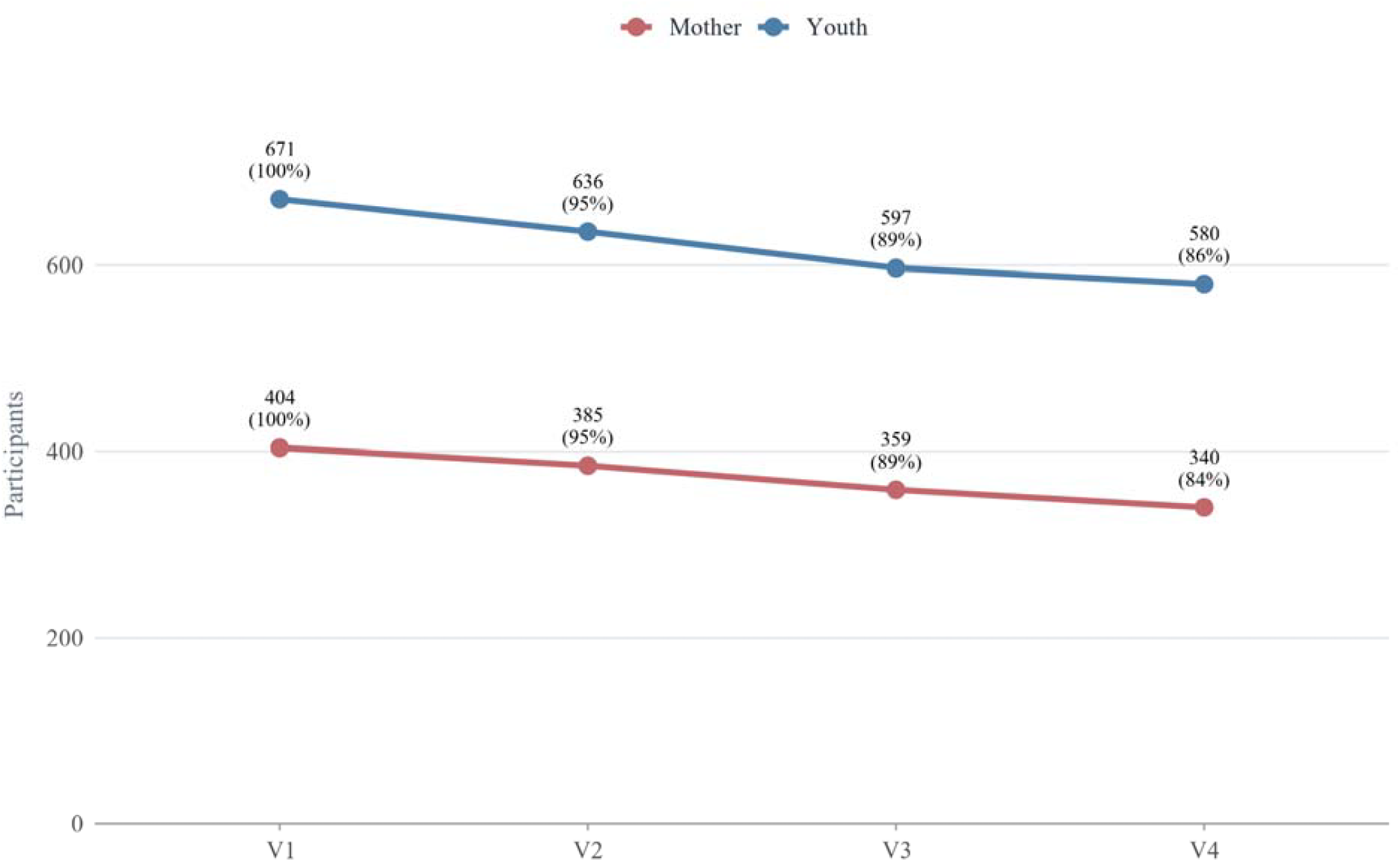
Cohort Retention Across Visits

**Figure 2:**
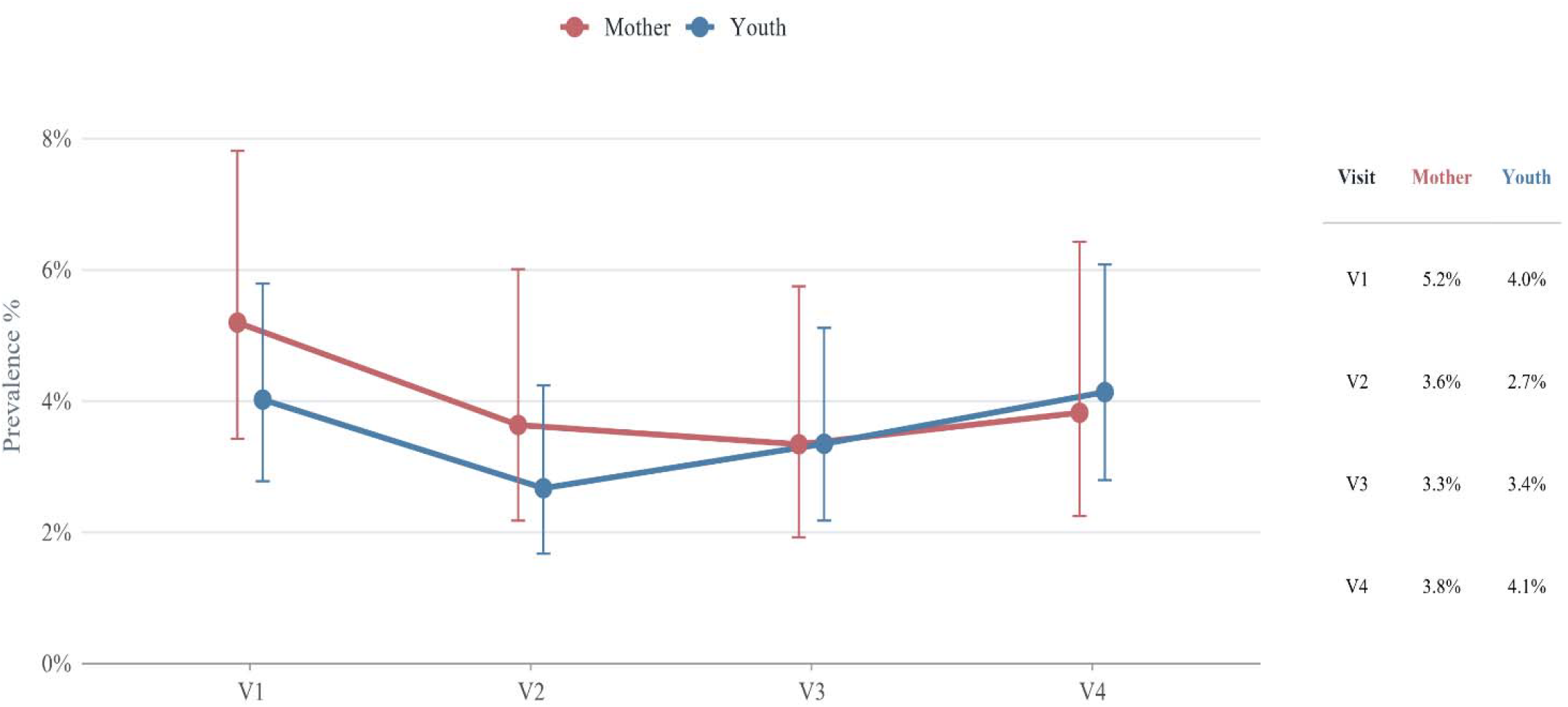
Oral HPV DNA Prevalence Across Visits

Despite this ongoing turnover, sustained infection was rare. Under the primary any-type definition, only 21 participants (1.9%) were positive at two or more visits (9 mothers [2.2%] and 12 youth [1.8%], representing 20.0% and 17.9% of ever-positive mothers and youth), and just 3 (one mother and two youth) were positive at all four visits; counts were stable across the type-specific and visit 2-4 definitions **(Supplementary Table 1)**. Transitions in oral HPV status between consecutive visits confirmed this transience, pooled across the three intervals, sustained positive-to-positive transitions comprised only 1% of all transitions in both cohorts, negative-to-negative transitions dominated (mothers 93.1%, youth 94.2%), and clearance and acquisition occurred at comparably low rates (mothers 3.3% and 2.6%; youth % and 2.4%). Persistence was concentrated among PLWH, the four-category classification differed by maternal HIV status (Fisher exact p=0.02) and by youth HIV exposure (p=0.04), and all nine persistent mothers and eight of twelve persistent youth were living with HIV.

HIV status was likewise the dominant correlate of detection and clearance. Among the 112 ever-positive participants, 73 cleared the infection, 37 remained positive at their final observed visit, and 2 were lost to follow-up while still positive and were censored. In modified-Poisson models, HIV was associated with higher visit-level oral HPV detection, with larger associations among mothers (high-risk PR 3.46, 95% CI 1.2-10.03; any-HPV PR 4.16, 1.77-9.74) than youth (1.91, 1.04-3.51; and 1.67, 0.98-2.83) **(Figure 3)**; the generation × HIV interaction was 0.55 (0.16-1.92), consistent with a weaker HIV-HPV association among youth, though imprecise. In a participant-level two-state Markov model fit to all observed person-intervals (71 acquisition and 80 clearance transitions, the clearance transitions exceeding the 73 participant-level clearances because participants with recurrent infection contribute more than one), clearance was significantly slower in PLWH than in those without HIV (clearance HR 0.42, 95% CI 0.22-0.77), whereas acquisition did not differ by HIV status (incidence HR 0.77, 95% CI 0.39-1.50) **(Supplementary Table 2)**.

**Figure 3:**
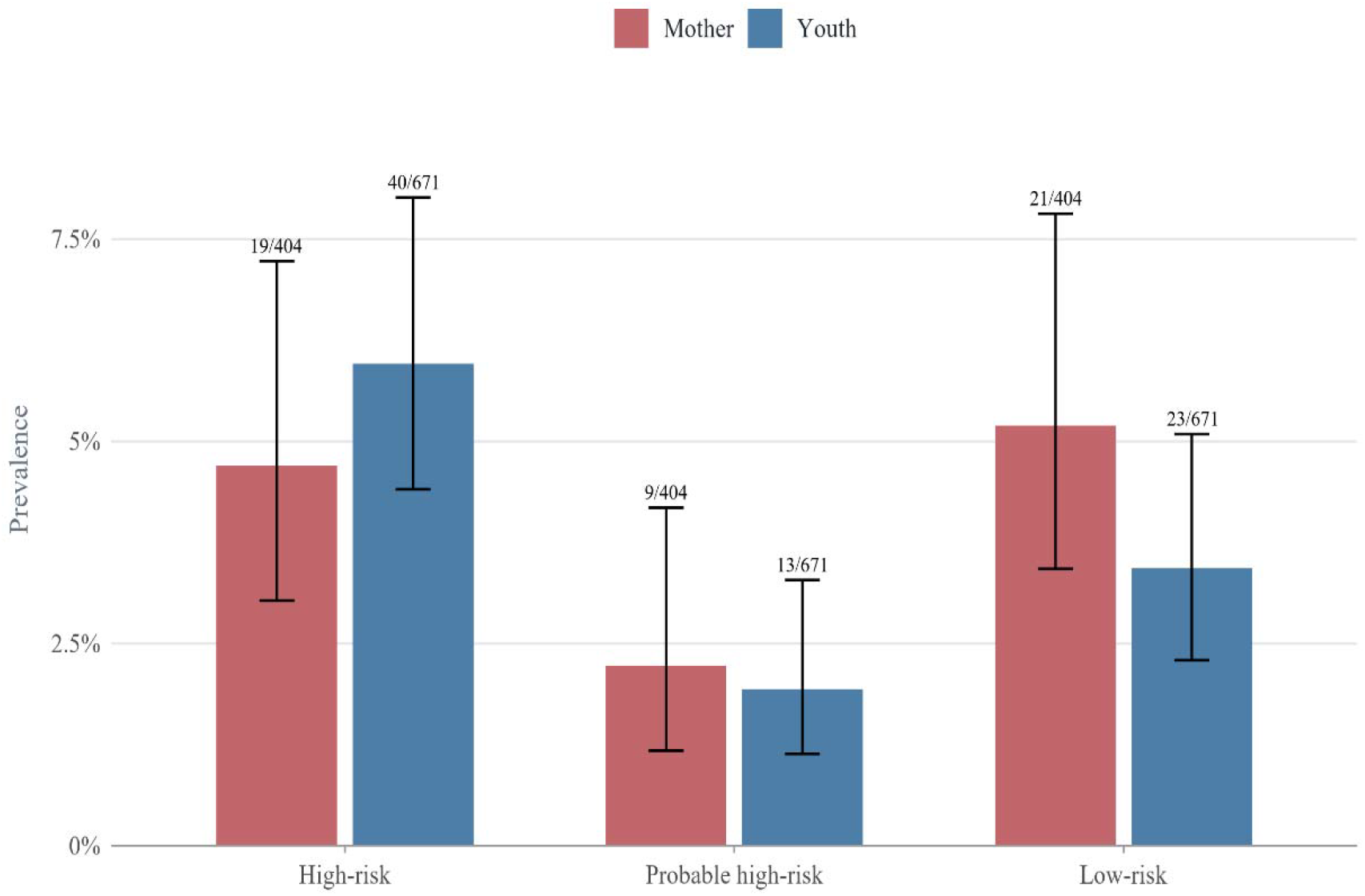
Oral HPV Prevalence by Genotype Risk Tier

The circulating genotypes carried a clear prevention signal. High-risk genotypes were detected in 53 participants (4.9%), most frequently HPV16 (n=14, 1.3%) and HPV39 (n=13, 1.2%), followed by HPV53 (n=10), HPV6 (n=9), HPV40 and HPV66 (n=8 each), and HPV35 (n=7) **(Figure 3)**. Only 35 of the 112 ever-positive participants (31.2%) carried a Gardasil-9-preventable genotype, the remaining 68.8% carrying non-vaccine genotypes exclusively; the pattern was starkest among the three participants positive at all four visits, where both youth carried a single high-risk, non-vaccine genotype throughout (HPV59 and HPV39) and the mother carried low-risk HPV70 and HPV53 **(Figure 4)**. Beyond HIV status, persistent and transient participants were broadly similar in age, education, oral-health indices, and behavioral exposures. Finally, genotype sharing within mother-youth dyads exceeded chance, among the 105 informative dyads (≥1 member ever-positive), 12 (11.4%) had both members positive and 3 of these (25.0%) shared an identical genotype, exceeding the 0.41 concordant dyads expected under 10,000 random re-pairings (permutation p=0.009) **(Figure 5)**.

**Figure 4:**
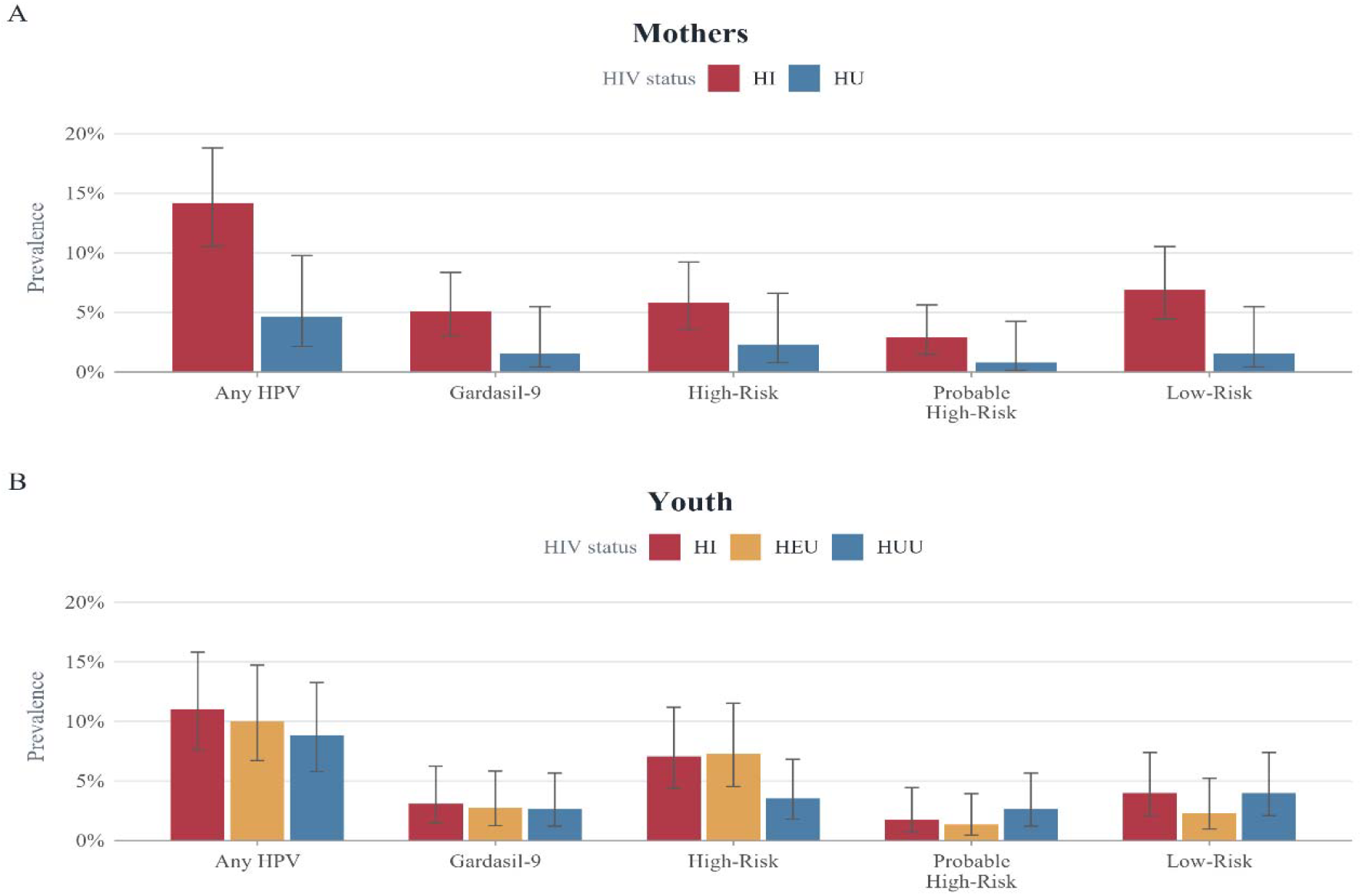
Oral HPV Prevalence by HIV Status. 5A. Mothers. 5B. Youth

**Figure 5:**
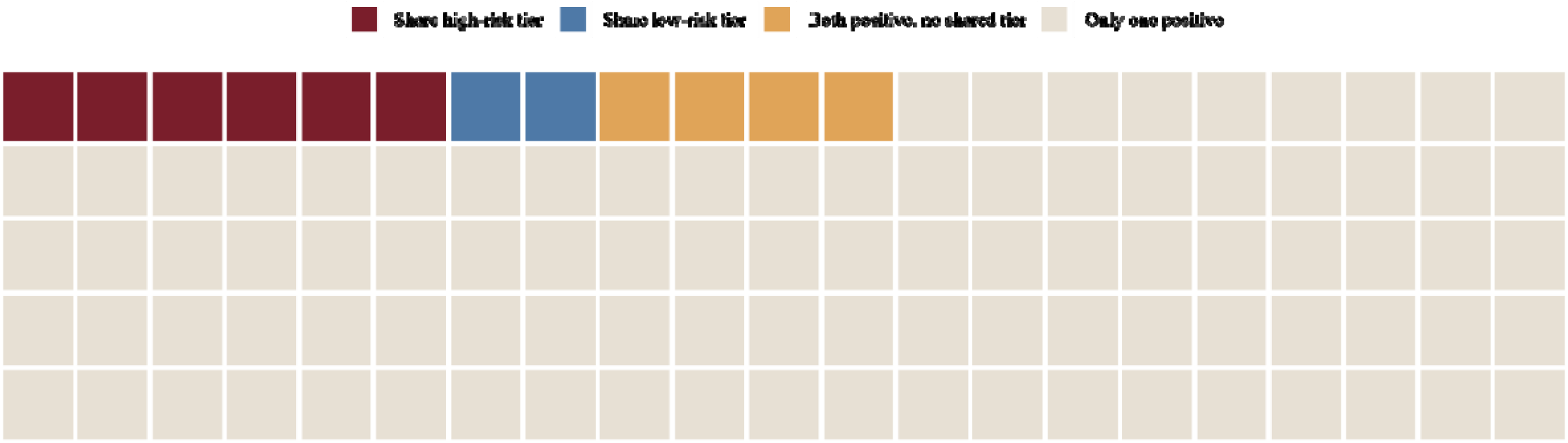
Risk Tier Sharing Across 105 Dyads that were positive at atleast one out of four visits.

## Discussion

To our knowledge, this prospective study offers one of the first longitudinal portraits of oral HPV natural history in a sub-Saharan African mother-youth population, and its central message is one of transience. Oral HPV was readily detected but rarely durable; most infections were acquired and cleared within the span of follow-up, and sustained persistence was the exception rather than the rule. Two features distinguished the small group in whom infection did persist. First, persistence clustered among PLWH, and second, the persisting genotypes were overwhelmingly high-risk types that current vaccines do not cover. Although the persistent fraction was small and these observations are necessarily descriptive, each aligns with evidence from better-studied anatomic sites and populations, and together they sharpen two priorities for this setting; surveillance focused on PLWH, and vaccines matched to the genotypes that actually circulate.

Our overall detection frequency of 10.4% sits at the upper end of the range reported for general adult populations, where pooled oral HPV prevalence is approximately 4.9% overall and high-risk prevalence approximately 2.6%^28^, and where standardized multinational sampling has reported any-type prevalence near 7.4%.^29^ The higher figure here is consistent with a cohort enriched for HIV infection and exposure, since people living with HIV carry oral HPV more often than HIV-negative individuals^30^, and recent sub-Saharan African cohorts report elevated oral HPV prevalence in HIV-affected adults.^31–33^ More important than the point prevalence, however, is the natural history our repeated sampling revealed. The predominance of clearance and the rarity of sustained oral HPV positivity in our cohort are consistent with longitudinal studies of men reporting low oral HPV incidence and rapid near-complete clearance over one to two years^34–36^, as well as studies of HIV-affected cohorts, in which most oncogenic oral infections cleared within a median of less than two years.^7^ This transience is the expected consequence of effective mucosal immune clearance of most incident HPV infections^37^, and it is the reason that persistence, rather than one-time positivity, is the epidemiologically informative endpoint.

Persistence was correspondingly uncommon, and this too is concordant with the literature. In the HPV Infections in Men (HIM) cohort, most oral infections cleared and long-term persistence of even HPV16 was rare^38^; in a seven-year HIV-affected cohort, 97% of incident and 71% of prevalent oral infections had cleared by the end of follow-up.^7^ Only three participants were positive at all four visits, and that estimates were stable whether we required the same genotype or restricted to later visits, indicates that the small, persistent fraction is a genuine feature of the data rather than an artifact of any single definition. The one consistent correlate of persistence was HIV status, all nine persistent mothers and two of three persistent youth living with HIV. This is the oral-mucosal counterpart of a well-established pattern at other sites, where HIV-related immunosuppression is associated with higher acquisition, slower clearance, and greater persistence of HPV, with lower CD4 counts and unsuppressed viral load independently predicting persistent infection^15,30,39^, and where women living with HIV carry a higher prevalence of high-risk and multiple-type infection.^10,11^ Because oral HPV16 viral load itself predicts long-term persistence in HIV-affected individuals^16^, and because high-risk oral HPV incidence is several-fold higher in men with HIV^17^, the concentration of persistence among HIV-infected participants in our cohort is mechanistically plausible even where the absolute counts are small.

The genotype distribution carries the clearest prevention message. HPV16 was the single most frequently detected type, consistent with its global dominance in both oral infection and HPV-attributable oropharyngeal cancer^3,7^, but it was closely followed by HPV39. The overall spectrum was strikingly weighted toward high-risk, non-vaccine types including HPV35 with both super-persistent youth carrying a single high-risk, non-vaccine genotype (HPV59 and HPV39) at every visit. This non-16 predominance echoes reports from other African HIV-affected oral cohorts, in which atypical, non-16 high-risk genotype distributions are common^31^, and it mirrors the cervix, where pooled surveys have long shown that HPV-positive Nigerian and sub-Saharan African women are relatively less likely to carry HPV16 and more likely to carry HPV35, 45, 52, 56, and 58.^8^ HPV35 is of particular regional importance as it is disproportionately common across sub-Saharan and West Africa, and a distinct A4 sub lineage associated with African ancestry confers elevated cervical carcinogenic risk.^9,40,41^ The practical consequence is quantified by our coverage estimate, and only 31.2% of ever-positive participants carried a Gardasil-9 preventable type, leaving more than two-thirds carrying non-vaccine types exclusively. Comprehensive cervical genotyping in the region reaches the same conclusion that the nonavalent vaccine, while highly beneficial, leaves a substantial residual fraction attributable chiefly to HPV35.^21,42^ This gap is widened in Nigeria specifically, where the national program introduced the single-dose quadrivalent vaccine (HPV6/11/16/18) in 2023^23^, a formulation that covers fewer high-risk types than the nonavalent vaccine and none of the non-16/18 high-risk types prominent locally. Single-dose delivery is itself well justified: randomized data from Kenya demonstrate high and durable efficacy of a single dose against persistent oncogenic infection^43,44^, immunobridging supports its use in the 9-14-year target age^45^, and WHO now endorses a one- or two-dose schedule.^46^ The limitation is one of valency and geography, not of dose, and our data reinforce the case for geographically informed, higher-valency formulations that include HPV35.^42^

This study has several strengths, including its longitudinal design with four visits in an under-studied population, a broad 28-genotype assay run under an identical protocol at two laboratories, the paired mother-youth structure, and high retention (85.6% of participants completing all four visits). Its limitations are equally clear. The number of persistent infections was small, so all persistence- and type-specific analyses are descriptive and hypothesis-generating. The study was powered for the cohort’s longitudinal incidence endpoints rather than for these sparse outcomes.^24,25^ Although HIV status was the consistent correlate of persistence, the small number of persistent participants precluded stable adjusted modeling of CD4 count, viral load, or antiretroviral therapy (ART) duration, so the mechanistic HIV story could not be quantified here. Additionally, oral rinse sampling captures oral but not specifically oropharyngeal or tonsillar epithelium, the site most relevant to HPV-associated oropharyngeal cancer.^47^ Genotype concordance cannot establish direction of transmission, and self-reported behavioral data are subject to reporting bias. Finally, the cohort is drawn from a single teaching hospital, which may limit generalizability.

In conclusion, oral HPV in this Nigerian mother-youth cohort was common but transient, with persistence confined to a small minority in whom HIV infection was the consistent feature. The genotypes that circulated and persisted were dominated by high-risk, non-vaccine types, HPV35 and HPV39, leaving more than two-thirds of infections outside nonavalent-vaccine coverage and a larger fraction outside the quadrivalent vaccine now used nationally in Nigeria. Even as a descriptive study, these findings argue that HIV-affected populations warrant particular attention in oral HPV surveillance, and that the durable reduction of HPV-associated disease in West Africa will depend on vaccine formulations designed to match the genotypes that circulate in the region.

## Availability of data and materials

The deidentified datasets generated and analyzed during this study are not publicly available due to ethical and regulatory restrictions associated with human participant data, including protections for sensitive health information. Data may be made available upon reasonable request to the corresponding author, contingent on institutional review board approval and completion of appropriate data use agreements.

## Authors’ contributions

a. AK contributed to the analysis and interpretation and drafted and critically revised the manuscript.
b. OP contributed to the analysis and interpretation and critically revised the manuscript.
c. EO contributed to the study conception, design, analysis, interpretation, and acquisition and critically revised the manuscript.
d. JC contributed to the analysis and critically revised the manuscript.
e. PA contributed to the study conception, design, analysis, and interpretation and critically revised the manuscript.
f. JL contributed to the analysis and interpretation and critically revised the manuscript.
g. NLI contributed to the study conception, design, analysis, and interpretation and critically revised the manuscript.
h. PK contributed to interpretation and critically revised the manuscript.
i. OO contributed to the study conception, design, analysis, and interpretation and critically revised the manuscript.
j. FEEU contributed to the study conception, design, analysis, and interpretation and critically revised the manuscript.
k. NFS contributed to the acquisition, analysis, and interpretation of the data and critically revised the manuscript.
l. YB contributed to the study conception, design, analysis, acquisition, and interpretation and critically revised the manuscript.
m. NOP contributed to the study conception, design, analysis, acquisition, and interpretation and critically revised the manuscript.
n. MOC contributed to the study conception, design, analysis, acquisition, and interpretation and critically revised the manuscript.

## Acknowledgements

The authors are grateful to the women, children/adolescents and families who made this study possible and to the dedicated study staff at the University of Benin Teaching Hospital and the Institute of Human Virology, Nigeria. Special thanks to all the HOMINY study team members, including Lydia Esena-Obaweiki, Promise Olumefun, Toluwalope Gbolahan, Isioma Mayor-Nwakwuribe, Alex Olanipekun, Amara Godwins, Modupe Kadiri, Daniel Oakhu, Matthew Imoe-Auweokha, Jessica Kubeyinje, Daniel Anonyai, Erika Juhlin, and Dr. Uwagboe Odigie, for their invaluable contributions to participant engagement and recruitment, as well as sample and data collection and analysis. The authors also appreciate the dedicated efforts of the study dentists - Drs. Nneka Chukwumah, Stanley Iyorzor, Oseriemen Akhigbemen, Timothy Ahworegba, Gbenga Ojuola, and Sunday Adesiyan—for their time and expertise in conducting clinical examinations during study visits. Many thanks to Jianhong Chen and the team at Inqaba Biotec West Africa for their support in laboratory management, sample processing, and HPV DNA detection and subtyping.

## Legends for Figures and Tables

Figure 1. Participant flow and retention across the four study visits.

Figure 2. Visit-specific oral HPV DNA prevalence across the four visits, by cohort.

Figure 3. Oral HPV prevalence by genotype risk tier. Bars show the percentage of participants positive for high-risk versus low-risk genotypes

Figure 4. Visit-specific oral HPV prevalence by HIV status, shown separately for mothers (4A: HI vs HU) and youth (4B: HI, HEU, HUU).

Figure 5. Risk-tier sharing within mother-youth dyads.

