## Supplementary Tables for "Natural History Of Oral Human Papillomavirus In Nigerian Mothers And Youth With And Without HIV: A Prospective Cohort Study"

Supplementary Table 1: *Persistence under four analytic definitions, expressed as a percentage of ever-positive participants.*

| **Definition** | **Mothers, n (%)** | **Youth, n (%)** |
| --- | --- | --- |
| Any-type, V1–V4 | 9 (20.0%) | 12 (17.9%) |
| Any-type, V2–V4 | 8 (17.8%) | 9 (13.4%) |
| Type-specific, V1–V4 | 7 (15.6%) | 12 (17.9%) |
| Type-specific, V2–V4 | 7 (15.6%) | 9 (13.4%) |

Supplementary Table 2. *Markov Model*

| **Transition** | **Transitions observed, n** | **Rate per visit-interval (95% CI)** | **Hazard ratio (95% CI)** |
| --- | --- | --- | --- |
| Acquisition (HPV− → HPV+) | 71 | 0.057 (0.040-0.080) | 0.77 (0.39-1.50) |
| Clearance (HPV+ → HPV−) | 80 | 1.75 (1.27-2.39) | **0.42 (0.22-0.77)** |
